# Social Network Analysis of COVID-19 Transmission in Karnataka, India

**DOI:** 10.1101/2020.08.11.20172734

**Authors:** S. Saraswathi, A. Mukhopadhyay, H. Shah, T.S. Ranganath

## Abstract

We used social network analysis (SNA) to study the novel coronavirus (COVID-19) outbreak in Karnataka, India, and assess the potential of SNA as a tool for outbreak monitoring and control. We analyzed contact tracing data of 1147 Covid-19 positive cases (mean age 34.91 years, 61.99% aged 11–40, 742 males), anonymized and made public by the government. We used software tools Cytoscape and Gephi to create SNA graphics and determine network attributes of nodes (cases) and edges (directed links, determined by contact tracing, from source to target patients). Outdegree was 1–47 for 199 (17.35%) nodes, and betweenness 0.5–87 for 89 (7.76%) nodes. Men had higher mean outdegree and women, higher betweenness. Delhi was the exogenous source of 17.44% cases. Bangalore city had the highest caseload in the state (229, 20%), but comparatively low cluster formation. Thirty-four (2.96%) “super-spreaders” (outdegree≥5) caused 60% of the transmissions. Real-time social network visualization can allow healthcare administrators to flag evolving hotspots and pinpoint key actors in transmission. Prioritizing these areas and individuals for rigorous containment could help minimize resource outlay and potentially achieve a significant reduction in COVID-19 transmission.

## Introduction

Karnataka, a southern state of India, reported its first case of coronavirus disease (COVID-19) caused by the severe acute respiratory syndrome coronavirus 2 (SARS-CoV-2), on 9 March 2020. The state has had fewer cases than other Indian states and has deployed modern technology tools as part of its logistics and containment measures[1,2]. As of 17 May 2020, Karnataka had declared 1147 diagnosed cases, 38 deaths, and 18648 individuals under observation[3]. Among the 1147, there were 600 active cases, 509 who had recovered, and 37 who died due to COVID (a fatality rate of 3.2%); one person died by suicide after being diagnosed. Social network analysis (SNA) is a technique to study the configurations of social relations between individuals or other social units. Social network models can be used to measure variables that shape relationships between social actors, and the extent to which they affect health-related outcomes[4,5]. Researchers are exploring the application of SNA to various facets of the COVID-19 pandemic, such as the role of public figures in communication[6], and clustering patterns within the broader patient network[7].

The Karnataka healthcare task force has relied primarily on contact tracing to limit the spread of COVID-19. To aid in accelerating our understanding of the transmission characteristics of this novel virus, we applied SNA to the contact tracing data of COVID-19 patients from Karnataka State, with two main research objectives in mind: First, can SNA improve our understanding of the transmission patterns of SARS-CoV-2? Second, can SNA produce actionable findings that can help in timely control of the spread of this disease?

## Methods

We performed social network analysis (SNA) on the anonymized contact tracing data of 1147 SARS-CoV-2 positive patients, uploaded to the public domain by the state government of Karnataka[3]. Our analysis included all cases reported positive for COVID-19 from 9 March to 17 May 2020, spanning the period from detection of the first Covid-19 case in the state to the end of phase-3 of the preventive lockdown declared by the government. We collected anonymized contact tracing data from daily government bulletins, and tabulated and summarized relevant demographic details such as age, district of residence, and history of travel, using Microsoft Excel. We created nodes and links tables in Excel, with each node representing a patient, and each link (edge), a confirmed contact between a source and a target patient. We imported this dataset into Gephi version 0.9.2 and applied the following sequence of layout algorithms: YiFan Hu Proportional, Fruchterman Reingold, and ‘No Overlap,’ to achieve a visual representation in which the more connected nodes are placed centrally, and ones with lower connectivity are placed towards the periphery of the network[8].

We wanted to use the capabilities of the two leading software tools[9], Gephi and Cytoscape, and make use of the features missing in one but available in the other. The use of Gephi’s network analysis tools results in the data for nodes and edges being populated with additional attribute variables. These values, such as node betweenness and edge betweenness, can then be displayed as visual features of the network elements in other software tools such as Cytoscape[10]. We reformatted the data exported from Gephi to make it compatible with the data model acceptable to Cytoscape version 3.8.0, which we used to create network graphics highlighting pertinent demographic characteristics of the nodes. Layout algorithms provided in Cytoscape were applied in the following sequence: Compound Spring Embedder (CoSE) and yFiles Remove Overlap, followed by a few manual adjustments to improve clarity. Network attributes generated by Gephi were analyzed using MS Excel to explain relevant aspects of the network and its components.

## Important definitions

**Degree centrality** is a measure of the number of social connections or links that a node has. It is expressed as an integer or count[11]. The indegree of a node is the number of incoming links to it from source nodes and refers to the number of infectious patients who had confirmed contact with a given target patient. Outdegree is the number of links to target nodes from a source node and is a measure of the number of secondary cases infected by a given patient.

**Betweenness centrality** is a measure of the number of times a node appears on the shortest path between other nodes[12]. It reflects the role a patient plays in creating a bridge of infectious transmission between patients who would not have had direct contact with each other.

**Closeness centrality** is the average of the shortest path lengths from a node to every other node in the network. We used harmonic closeness to measure closeness centrality due to the presence of unconnected nodes[13].

**Edge betweenness** is the number of the shortest paths that go through an edge in a graph or network, with a high score indicative of a bridge-like connection between two parts of a network, crucial to transmission between many pairs of nodes[14].

**Clustering coefficient** measures the degree to which nodes in a graph tend to cluster together[15].

**Network density** is the number of existing ties between nodes, divided by the number of possible ties[16].

**Network diameter** is the shortest path between the two most distant nodes in a network[15].

**Mean path length** is the average of the shortest path lengths between all possible node pairs[15].

**Network component** is an island of interlinked nodes that are disconnected from other nodes of the network. Many networks consist of one large component, sometimes together with several smaller ones and singleton actors[5].

**Super-spreader** (operational definition): Any node with an outdegree ≥5 was considered a super-spreader. Individuals represented by these nodes would have infected five or more contacts.

## Results

Demography: We analyzed 1147 patients (742 males, 64.69%), aged 34.91 years on average, most of whom (711, 61.99%) belonged in the 11–40 years age range. Most deaths, however, occurred among older patients, with the highest mortality percentage (10/34, 29.41%) in patients aged over 70 years (Supplementary Figure S1).

Network parameters (Supplementary Table S1): We found 948 nodes with zero outdegree. The remaining 199 (17.35%) nodes had an outdegree range of 1–47 and were the source of infection to 657 targets through 706 links (edges). Among the target nodes, 36 had indegree >1 (range 2–5), implying more than one source. There were 490 nodes with zero indegree, of which 383 had zero outdegree. These were isolated nodes with degree centrality value zero. The range of betweenness centrality was 0.5–87 for 89 (7.76%) nodes. The network had 143 nodes with a harmonic closeness centrality (HCC) of one and 56 with HCC between zero and one. Our network density was 0.001, diameter was 4, and clustering coefficient was 0.004.

Men had a higher mean outdegree (0.628, M vs 0.593, F) and women, higher betweenness (0.573, F vs 0.412, M, Supplementary Table S2). The 95^th^ percentile values for outdegree and betweenness were 3 and 2, respectively. There were 77 (6.71% of 1147) nodes with outdegree ≥3, and combined, they accounted for 556 (78.75% of 706) edges. More than two-thirds of these (54, 70.13%) were men. The average HCC for the 77 nodes with outdegree≥3 was 0.887, compared to 0.161 for the entire network. Of the 59 nodes with betweenness ≥2, more than half (33, 55.93%) were men, though women had a higher mean betweenness overall.

We noted 34 super-spreaders with outdegree ≥5, with a cumulative outdegree of 410, and after deducting 17 duplicate edges for target nodes with indegree >1, they accounted for 393 (59.81%) of the 657 target cases.

The aggregate network graphic (Figure 1), created using Gephi, shows nodes representing patients, and components representing case clusters. The nodes are colored according to district and sized by outdegree, making the larger nodes represent individuals who infected a greater number of targets. Bangalore had the highest number of cases, followed by Belagavi, Kalaburagi, and Mysuru (Supplementary Table S3). The largest node is in Mysuru, denoting a patient who infected 47 target nodes, at the center of a major component. Transmission between districts was limited, occurring chiefly from Mysuru to Mandya, a geographically adjacent city. The network figure also has two large-sized gray nodes that represent two patients with outdegree 29 and 25, from districts Vijayapura and Uttara Kannada, respectively.

**Figure 1:**
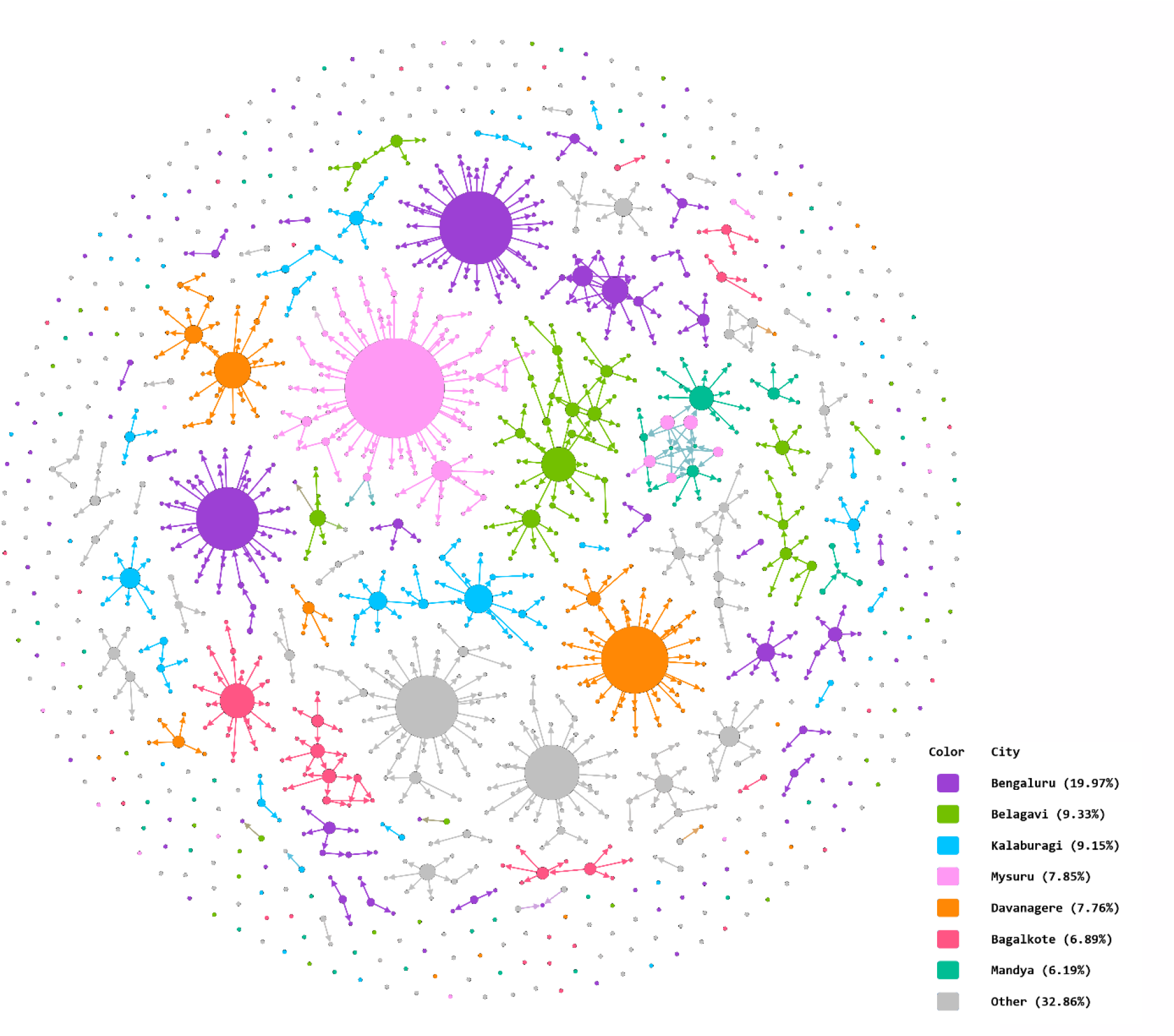
Aggregate Network Graphic Created in Gephi. Arrowheads indicate direction of transmission from source node to target node. Node size reflects outdegree. Edges inherit color from parent nodes.

The network contained 93 clusters of connected nodes (components), of which 37 components, made up of five or more nodes each, had more than half of all the nodes (613, 53.44%) and four-fifths of the links (611, 86.54%) concentrated within them (Figure 2).

**Figure 2:**
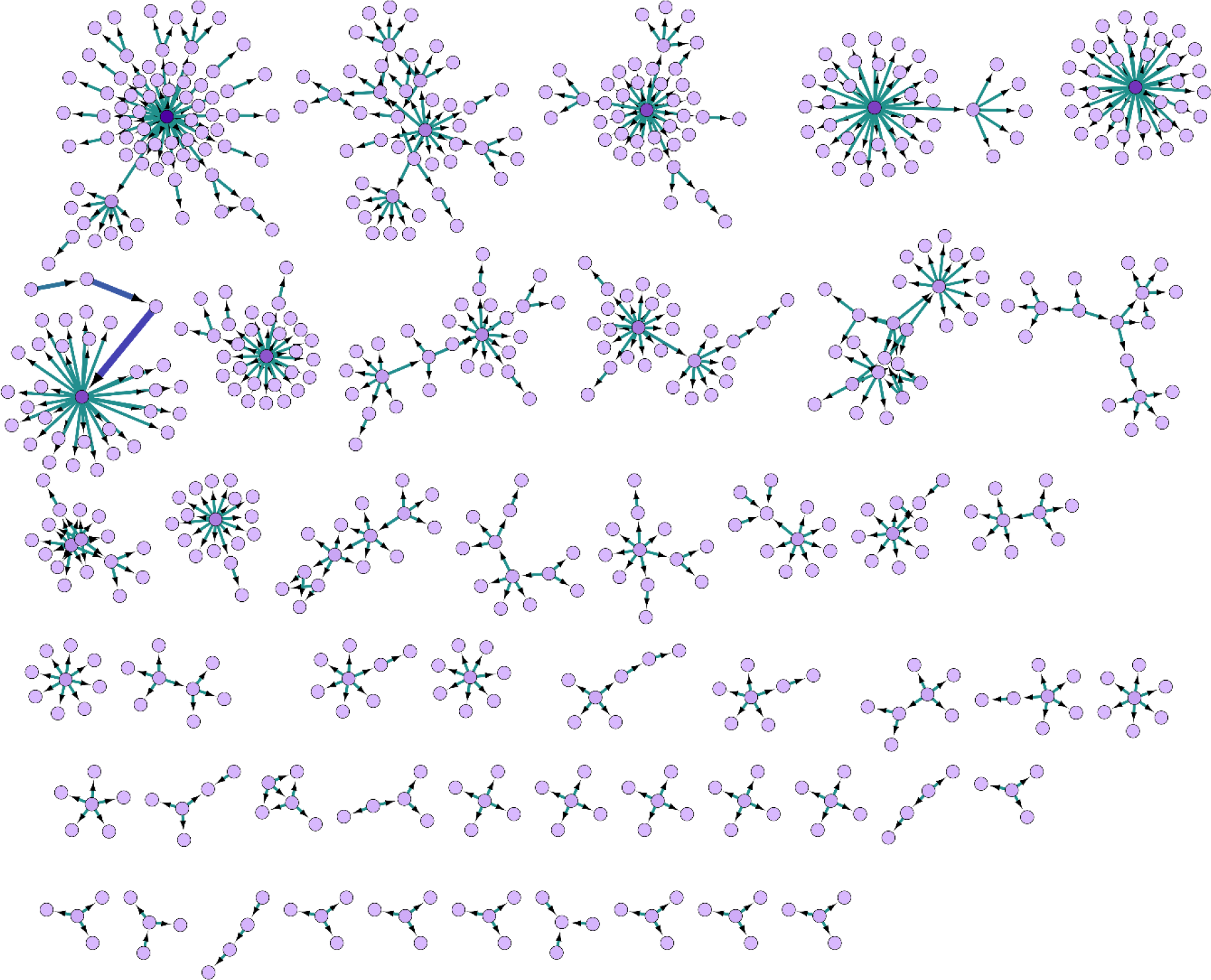
Major Network Components Organized by Size, Created in Cytoscape. Arrowheads indicate direction of transmission from source node to target node. The thickness and color intensity of edges reflect edge betweenness.

Figure 3 shows the age and sex distribution of cases in the network. Nodes are colored by age group and sized by outdegree. Figure 4 shows nodes colored by source of infection and sized by betweenness centrality. We have considered patients with a history of travel from Delhi in a separate category as their count was comparable to the combined number of travelers from all other states of India. It is noteworthy that travelers from abroad did not contribute to the formation of any major cluster.

**Figure 3:**
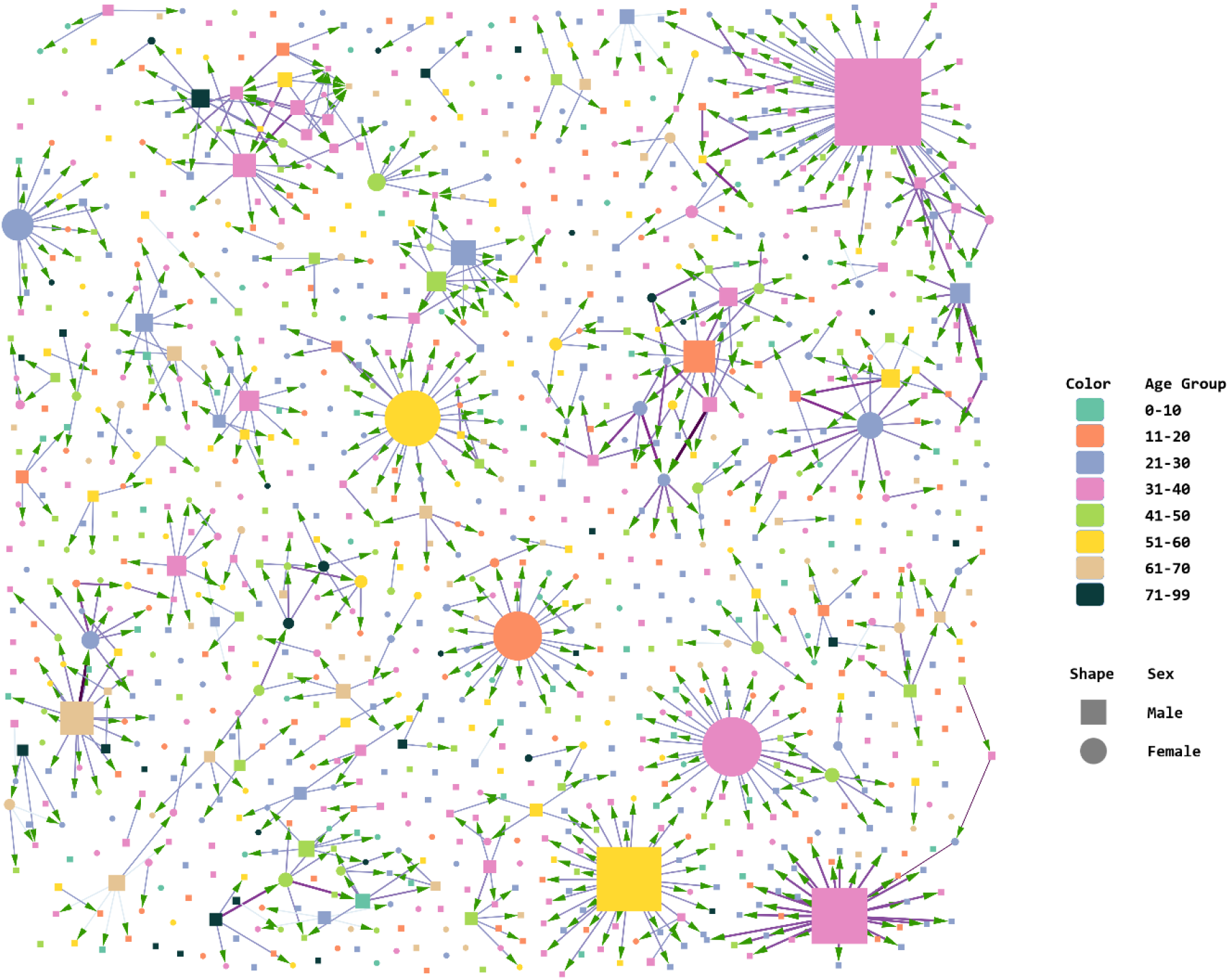
Age-Sex attributes of Nodes and Clusters, Created in Cytoscape. Node size indicates outdegree centrality. Arrowheads indicate direction of transmission from source node to target node. The thickness and color intensity of edges reflect edge betweenness.

**Figure 4:**
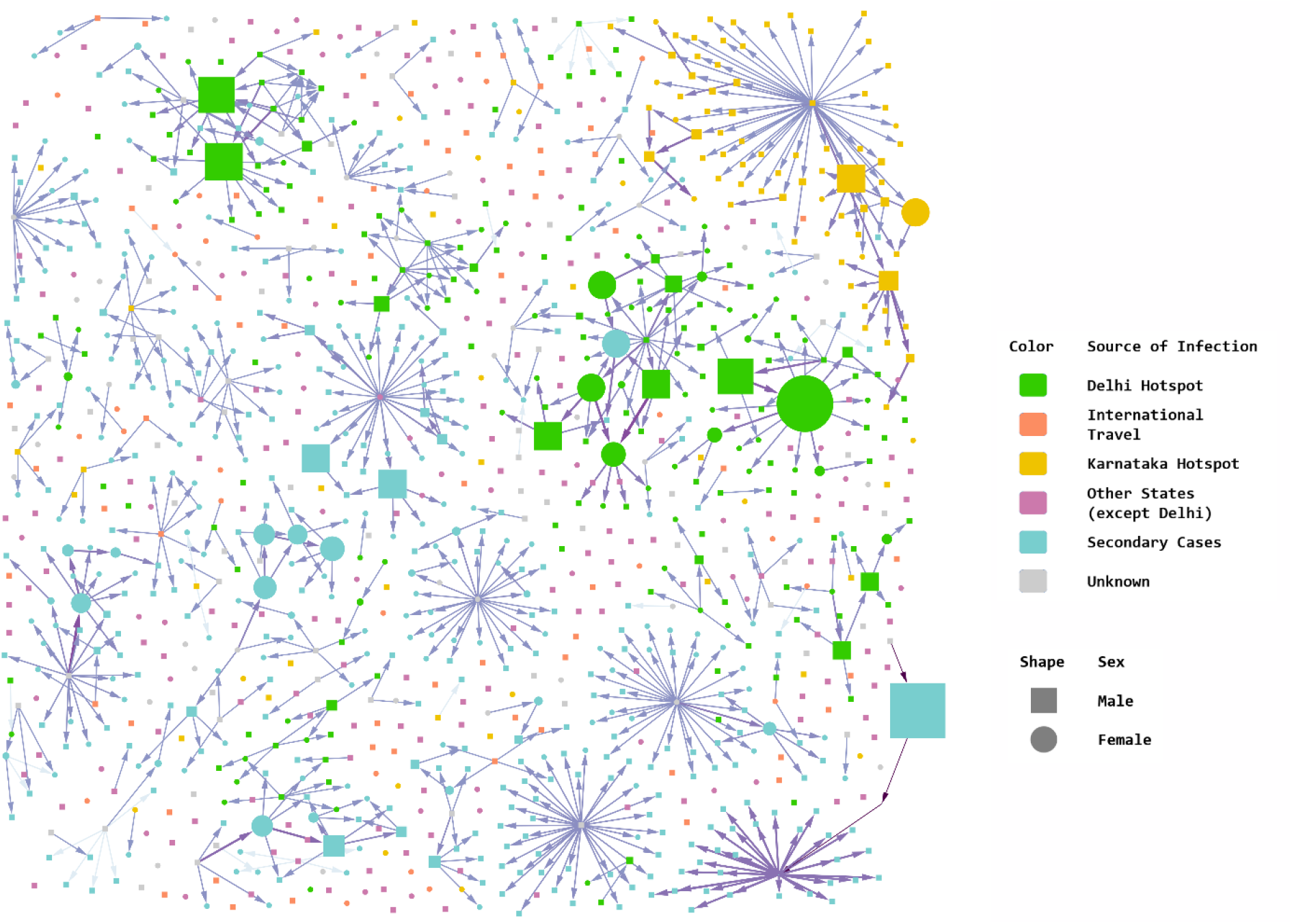
Network Analysis by Sources of Infection (Cytoscape) Node size indicates betweenness centrality. Arrowheads indicate direction of transmission from source node to target node. The thickness and color intensity of edges reflect edge betweenness.

Comparing figures 3 (nodes sized by outdegree) and 4 (nodes sized by betweenness), we find that in clusters with nodes that had multiple interconnections, relatively low outdegree, and high betweenness, the key nodes were females. This indicates that women played a significant bridging role. This differs from clusters with edges radiating from a central node with high outdegree and low betweenness, where typically, a young male was the nidus. The largest and second-largest components illustrate this difference in transmission (Figure 5). The largest component had 75 nodes and 76 links, and the second-largest component had 45 nodes and 50 links. The largest cluster originated in the district of Mysuru; its source node was a male with high outdegree who spread the infection to many contacts. However, secondary transmission from those contacts was limited. This cluster is star shaped. The second-largest component resembles a spiderweb with multiple interconnected nodes and many female actors. This cluster was in Belagavi, and its network density was nearly twice that of the largest cluster (.025 vs. .014), with a shorter average path length (1.314 vs. 1.321).

**Figure 5:**
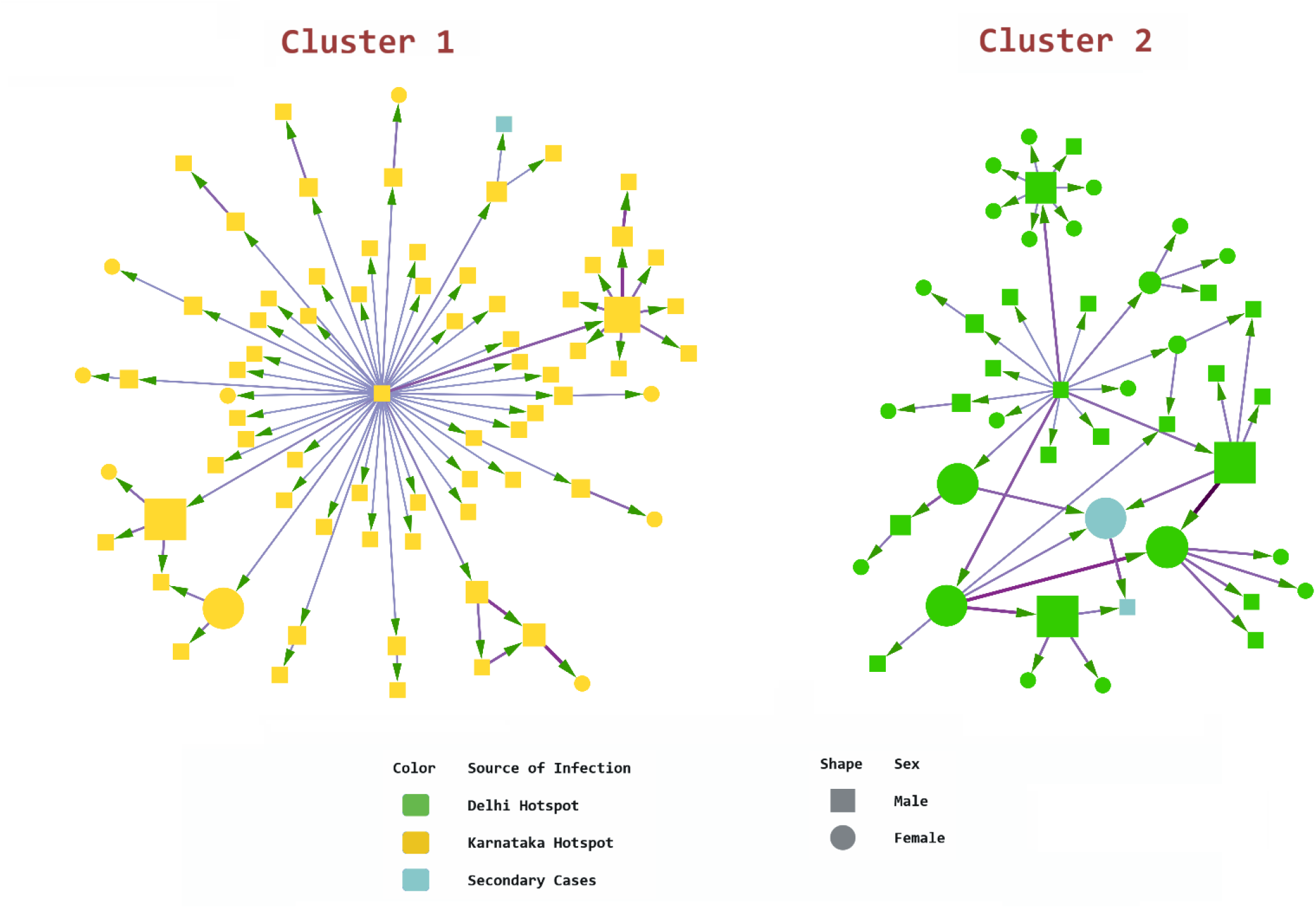
Comparing the Two Largest Components (Cytoscape) Node size indicates betweenness centrality. Arrowheads indicate direction of transmission from source node to target node. The thickness and color intensity of edges reflect edge betweenness.

## Discussion

Our study reveals that most cases of COVID-19 in Karnataka were young and middle-aged men. Deaths, however, occurred overwhelmingly among elderly patients. Close to one-third of those aged ≥60 years (35/112, 31.25%) were secondary cases who had contracted COVID-19 from younger contacts, and for another 25% (28/112), the source of infection was unknown with no history of travel to or from hotspots. It is plausible that these latter were also contacts of SARS-CoV-2 carriers. The age and sex profile of our study set matches nationwide surveillance data from India, reported by Abraham et al., with median age and age-distribution close to our sample, and a similar high attack rate in males[17].

Bangalore, the capital of Karnataka, is a densely populated metropolis, housing one-sixth of Karnataka’s population in one per cent of its area[18,19]. The city airport is a major transit point for domestic and international travelers. These factors may explain Bangalore’s relatively heavy burden (229/1147) of COVID-19 cases. Despite accounting for nearly a fifth of the state’s caseload, however, our network analyses (Figure 1 and Supplementary Tables S3 and S4) show that Bangalore did not have notably large clusters compared to other districts. Most of the cases detected here were isolated nodes, many of whom were returnees from abroad. Bangalore’s low transmission may be due to the disciplined observance of lockdown measures, and rigorous contact tracing and quarantine activities by its healthcare workforce[20,21].

The presence of two large nodes (where size denotes outdegree) in districts that had a minor contribution to the total caseload (Figure 1) points to the risk of cluster formation even in relatively unaffected areas if distancing measures are not followed scrupulously.

Shortly after the World Health Organization confirmed the novel Coronavirus as the cause of the outbreak in China[22], health authorities started precautionary screening at Bangalore’s international airport, and quarantining passengers arriving from areas of concern[23]. These early steps may explain why we found no major clusters originating from international travelers. Conversely, we noted several clusters comprised of people with a history of travel to the national capital, Delhi (Figure 4 and Supplementary Tables S3 and S4). By 19 April 2020, the entire city of Delhi had been declared a COVID-19 hotspot[24] in the wake of a mass religious gathering that was found to be linked to nearly a third of the country’s caseload earlier in the month[25]. Our second-largest cluster was traced to a patient who had a history of attending this religious gathering in Delhi. Clusters of cases that originated from Delhi tended to be closely interconnected, with women playing an active transmission role. This could reflect close community ties between these individuals, or residence in underprivileged areas where strict social distancing may not have been observed.

Most of the clusters in our network showed a man with high outdegree as the nidus. Women, on the other hand, played an important role in transmission by bridging multiple nodes within clusters, even though men outnumbered women in the 95^th^ percentile region of betweenness. Further study is warranted into the behavioral characteristics of men and women that drive these differences.

The low density of our network, the presence of 948 nodes with zero outdegree, and the fact that only 34 source cases had infected close to two-thirds of all target cases, indicate that community transmission was negligible. Bi et al. reported a similar transmission pattern from Shenzhen, China. In their cohort, 8.9% of the cases had caused 80% of all infections[26]. Network analysis of COVID-19 patients in Henan, China, by Wang et al.[7] revealed a similar non-uniform pattern of clustering (208/1105 patients in clusters) with a skewed distribution of patients in different cities. Their findings also indicate a strong correlation of confirmed cases with travel to Wuhan (the epicentre of the pandemic), which is concordant with our observation that a significant proportion (17.44%) of the Karnataka patients had traveled to Delhi.

### Limitations

Our SNA findings may not universally reflect field realities. Some findings such as eccentricity and mean path length are theoretical constructs computed by software algorithms, but in practice, these metrics remain indeterminate as our network had very few inter-district connections and many isolated nodes and components. Our dataset included many patients with contact tracing still under investigation at the time of analysis. We were not able to analyze the role of type and duration of contact, as the data for these were not available for many patients.

## Conclusion

Our conventional analysis indicates that senior citizens, due to their high mortality risk with COVID-19, should be advised strict social distancing, and older patients from rural or underserved areas should be preemptively transferred to tertiary centres with intensive care facilities. Our network analysis suggests that geographical, demographical, and community characteristics could be influencing the spread of COVID-19. Gender influences cluster morphology, with men seeding the clusters and women propagating them. Furthermore, our SNA highlights the need to maintain an accurate database with ongoing recording of contact tracing data using a uniform format. Tools for real-time visualization of social networks can provide actionable information on the evolution and spread of the disease. Such methods could aid local government bodies in formulating control measures tailored to network characteristics of each locality. Social network analysis can flag evolving networks with high densities and pinpoint nodes with high outdegree, betweenness, and closeness scores, which imply an active role in the transmission and bridging of infection. Public health authorities could prioritize these clusters and individuals for rigorous containment, which could help minimize resource outlay and potentially significantly reduce the spread of COVID-19.

## Data Availability

We used anonymized secondary data available in the public domain, from a government website, the copyright policy of which indicates that the data may be used for publication subject to proper acknowledgment. Archived bulletins will be made available on reasonable request from the principal author.

https://covid19.karnataka.gov.in/govt_bulletin/en#

## Author Contributions

S. Saraswathi: Conceptualization, Study Design, Data Collection

A. Mukhopadhyay and H. Shah: Data Analysis

All authors: Writing, Editing, Review and Final Approval of Manuscript

## Disclaimer, Funding, and Conflict of Interest

We used anonymized secondary data available in the public domain, from a government website, the copyright policy of which indicates that the data may be used for publication subject to proper acknowledgment. We received no financial support for this study. We have no conflict of interest to declare.

## Epidemiology and Infection

### Supplementary Material

**Figure S1:**
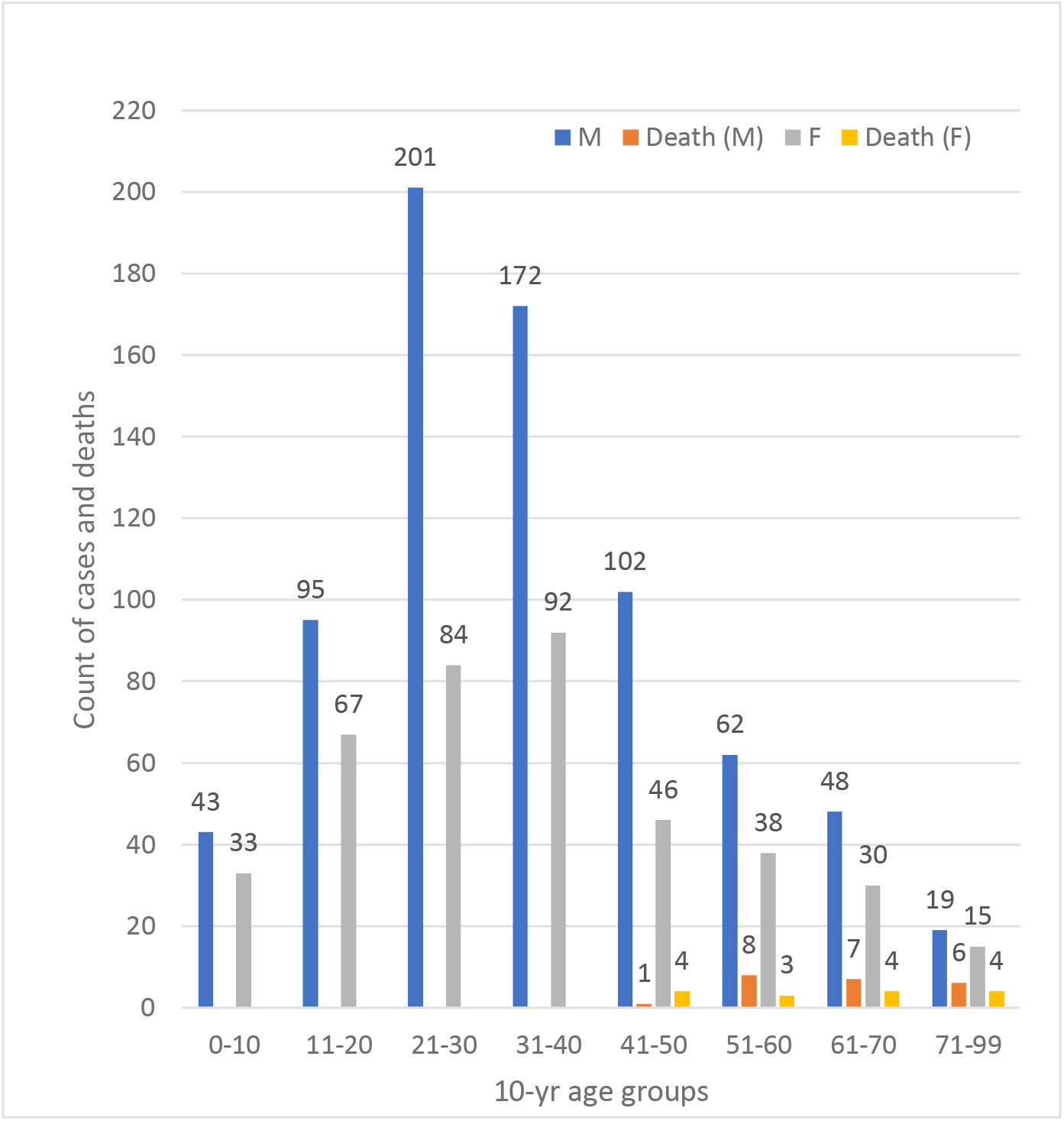
Age-Sex Distribution of Cases and Deaths

**Figure S2a:**
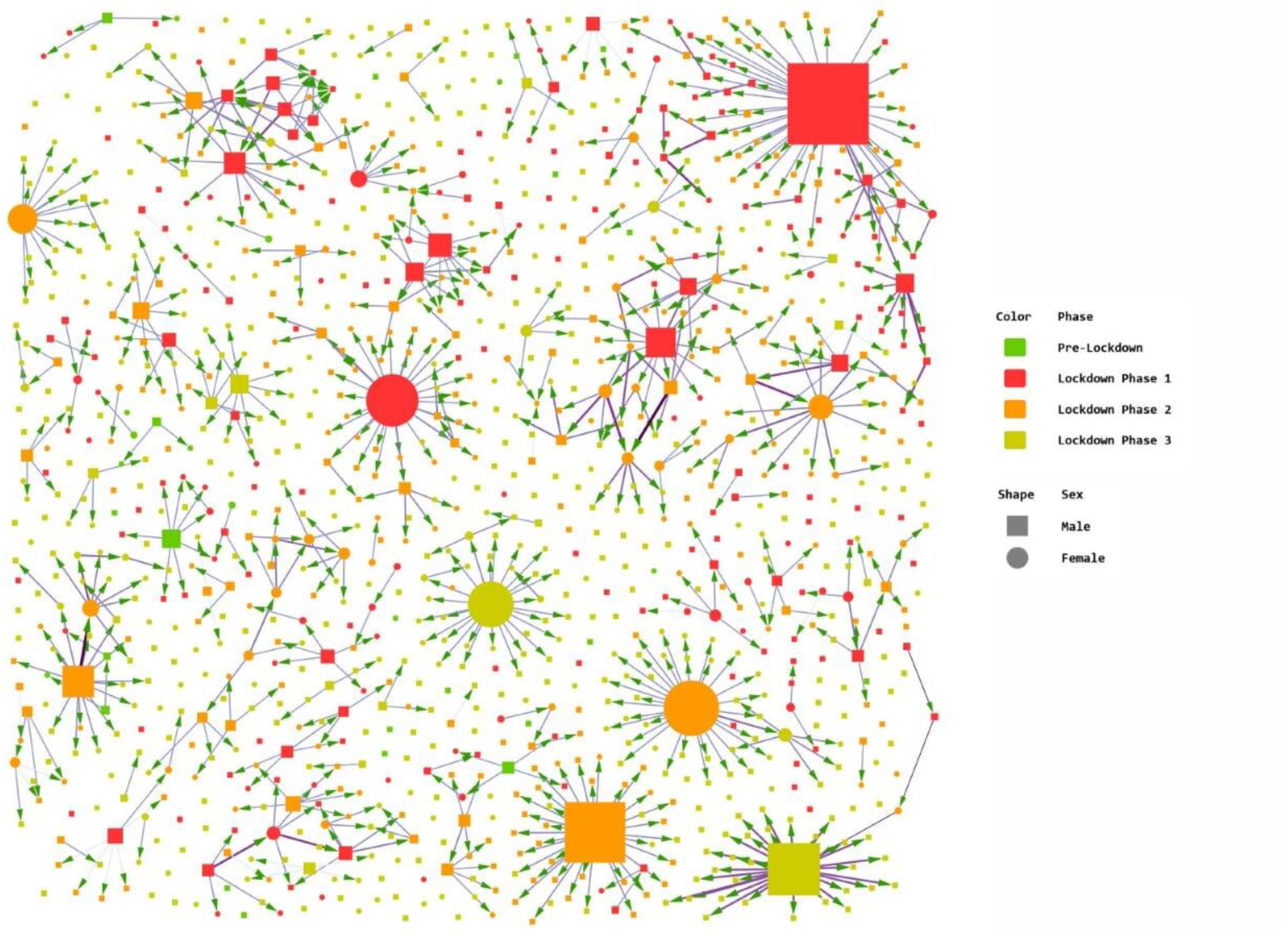
Network Analysis by Phases of Lockdown (Cytoscape) This figure shows the incidence of clusters during each phase of the preventive lockdown implemented by the government. Phase 1, with the most stringent curbs on travel and socialization, was from March 24 to April 14. The second phase spanned 19 days from April 15 to May 3, and the third phase was from May 4 to May 17. Several clusters evolved during the first lockdown phase as those infected in the pre-lockdown period turned symptomatic and tested positive. Many of these clusters comprised returnees from Delhi and their contacts.

**Figure S2b:**
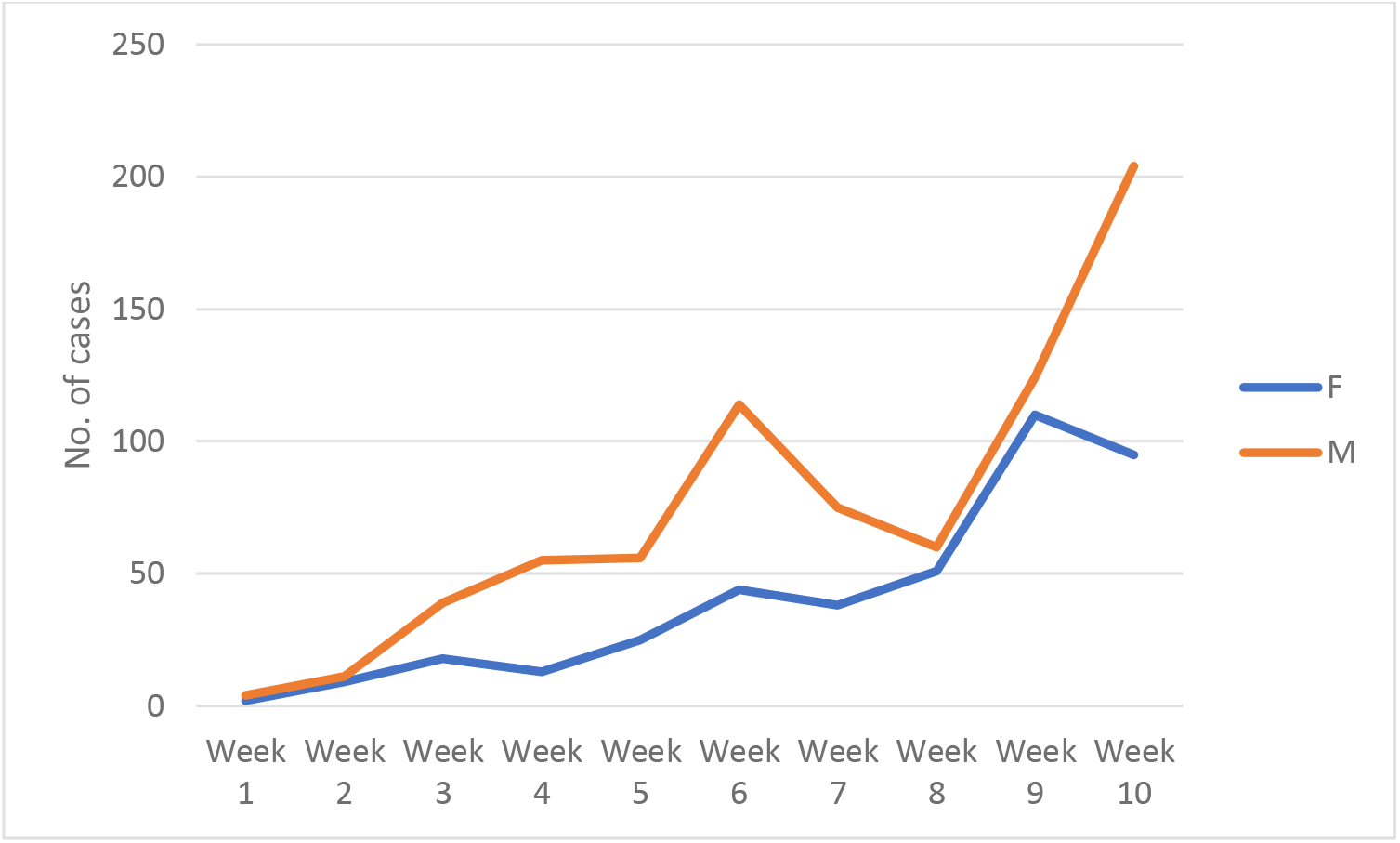
Trend of Cases (March 9 to May 17, 2020) This graph shows the number of cases detected every week. Week 1 begins on 9 March and week 10 ends on 17 May 2020. Cases spiked in the second half of April and continued to rise as lockdown regulations were relaxed and migrant workers returned from other states. However, these were mostly isolated nodes with few instances of cluster formation (Figure S2a), probably due to the effective implementation of quarantine measures.

**Table S1:**
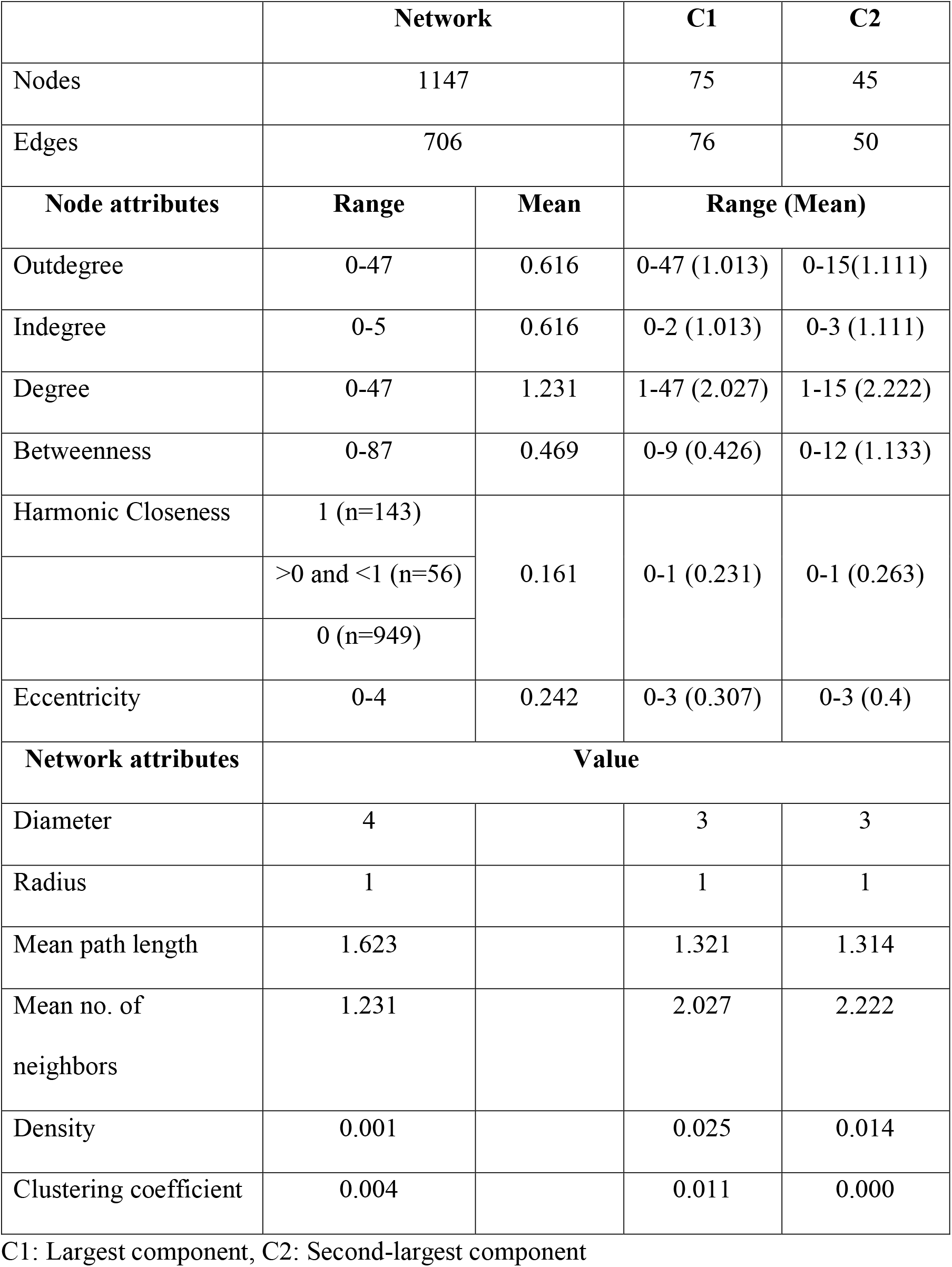
Network Parameters

**Table S2:** Mean Outdegree and Betweenness by Sex and Age-Group

| **Mean outdegree** | **F** | **M** | **Total** |
| --- | --- | --- | --- |
| 000-10 | 0.000 | 0.140 | 0.079 |
| 011-20 | 0.463 | 0.368 | 0.407 |
| 021-30 | 0.655 | 0.289 | 0.396 |
| 031-40 | 0.500 | 0.994 | 0.822 |
| 041-50 | 0.870 | 0.480 | 0.601 |
| 051-60 | 1.158 | 1.145 | 1.150 |
| 061-70 | 0.467 | 1.083 | 0.846 |
| 071-99 | 0.667 | 1.263 | 1.000 |
| **Total** | **0.593** | **0.628** | **0.616** |
| **Mean betweenness** | **F** | **M** | **Total** |
| 000-10 | 0.000 | 0.233 | 0.132 |
| 011-20 | 0.179 | 0.242 | 0.216 |
| 021-30 | 1.561 | 0.095 | 0.527 |
| 031-40 | 0.114 | 1.159 | 0.795 |
| 041-50 | 0.783 | 0.216 | 0.392 |
| 051-60 | 0.474 | 0.210 | 0.310 |
| 061-70 | 0.033 | 0.406 | 0.263 |
| 071-99 | 1.572 | 0.000 | 0.694 |
| **Total** | **0.573** | **0.412** | **0.469** |

**Table S3:** District-wise Caseload, and Sources of Infection

| **District** | **F** | **M** | **Total** | **%** |
| --- | --- | --- | --- | --- |
| Bengaluru | 75 | 154 | 229 | 19.97 |
| Belagavi | 46 | 61 | 107 | 9.33 |
| Kalaburagi | 45 | 60 | 105 | 9.15 |
| Mysuru | 11 | 79 | 90 | 7.85 |
| Davanagere | 37 | 52 | 89 | 7.76 |
| Bagalkote | 27 | 52 | 79 | 6.89 |
| Mandya | 24 | 47 | 71 | 6.19 |
| Others | 140 | 237 | 377 | 32.87 |
| Total | 405 | 742 | 1147 | 100.00 |
| **Source type** | **F** | **M** | **Total** | **%** |
| International travel | 24 | 66 | 90 | 7.85 |
| Domestic travel | 59 | 150 | 209 | 18.22 |
| Delhi hotspot | 70 | 130 | 200 | 17.44 |
| Karnataka hotspot | 23 | 90 | 113 | 9.85 |
| Secondary cases | 193 | 240 | 433 | 37.75 |
| Unknown | 36 | 66 | 102 | 8.89 |
| Total | 405 | 742 | 1147 | 100 |

**Table S4:** Distribution of the 37 Largest Clusters by District and Source of Infection

| Source<br>District | Unknown | Karnataka<br>hotspot | Delhi<br>hotspot | Domestic<br>travel | International<br>travel | Total |
| --- | --- | --- | --- | --- | --- | --- |
| Bengaluru | 3 | 2 | 2 |  | 1 | 8 |
| Belagavi |  |  | 5 |  |  | 5 |
| Kalaburagi | 3 |  | 1 | 1 |  | 5 |
| Mysuru |  | 1 |  |  |  | 1 |
| Davanagere | 5 |  |  |  |  | 5 |
| Bagalkote | 3 |  |  |  |  | 3 |
| Mandya |  |  | 1 |  |  | 1 |
| Others | 5 |  | 3 | 1 |  | 9 |
| Total | 19 | 3 | 12 | 2 | 1 | 37 |

Cell values indicate count of clusters.
Bengaluru, the major transit point for international passengers, had only one cluster traced to a returnee from abroad. It is notable that 11 of the 37 clusters (29.73%) originated in Delhi. All the Belagavi clusters (including the 2<sup>nd</sup> largest cluster with 45 nodes) were traced to travelers from Delhi.

